# Cytokine storm polymorphisms in nonvaccinated COVID-19 patients

**DOI:** 10.1101/2025.03.14.25323971

**Authors:** Laura E. Martínez-Gómez, Carla Isabel Oropeza-Vélez, Maylin Almonte-Becerril, Leslie Chavez-Galan, Carlos Martinez-Armenta, Rosa P. Vidal-Vázquez, Juan P. Ramírez-Hinojosa, Paola Vázquez-Cárdenas, Diana Gómez-Martín, Gilberto Vargas-Alarcón, José M. Rodríguez-Pérez, Lucero A Ramón-Luing, Julio Flores-Gonzalez, José G. Carrasco, Mónica M. Mata-Miranda, Gustavo J. Vázquez-Zapién, Adriana Martínez-Cuazitl, NM Parra-Torres, Felipe de J. Martínez-Ruiz, Dulce M. Zayago-Angeles, Ma. Luisa Ordoñez-Sánchez, Yayoi Segura-Kato, Carlos Suarez-Ahedo, Jessel Olea-Torres, Brígida Herrera-López, Carlos Pineda, Gabriela A. Martínez-Nava, Alberto G. López-Reyes

**Affiliations:** Laboratorio de Gerociencias, Dirección General, Departamento de Reconstrucción Articular. Instituto Nacional de Rehabilitación Luis Guillermo Ibarra Ibarra, Secretaría de Salud; Mexico City, México; Servicio de Medicina Interna de Hospital Petróleos Mexicanos PEMEX; Mexico City, México; Dirección Ejecutiva de Investigación y Estudios Avanzados (DEIEA), Universidad de la Salud (UNISA); Laboratory of Integrative Immunology, Instituto Nacional de Enfermedades Respiratorias Ismael Cosio Villegas, Mexico City 14080, Mexico; Centro de Innovación Médica Aplicada, Hospital General Dr. Manuel Gea González; Mexico City, Mexico; Department of Immunology and Rheumatology, Instituto Nacional de Ciencias Médicas y Nutrición Salvador Zubirán, Secretaría de Salud; Mexico City, Mexico; Departamento de Biología Molecular y Endocrinología, Instituto Nacional de Cardiología Ignacio Chávez; Mexico City, Mexico; Laboratorio de Biología Celular y Tisular, Laboratorio de Embriología, Escuela Médico Militar, Universidad del Ejército y Fuerza Aérea; Mexico City, Mexico; Nuevo Hospital General Delegación Regional Sur de la Ciudad de México ISSSTE; Mexico City, Mexico; Unidad de Biología Molecular y Medicina Genómica, Instituto Nacional de Ciencias Médicas y Nutrición; Salvador, Zubirán

**Keywords:** COVID-19, IL-6, IL-10, CCL-2, Polymorphism, SARS-CoV-2

## Abstract

Cytokines and chemokines are essential for establishing an appropriate immune response to severe acute respiratory syndrome coronavirus-2 (SARS-CoV-2). Variations in the genes encoding cytokines and chemokines strongly influence the immune response to pathogenic challenges and disease outcomes. This study was carried out to determine the associations of polymorphisms in the *TNF*-α*, IL-6*, *IL-8, IL-10*, and *CCL5* genes with COVID-19 severity. A total of 627 unvaccinated COVID-19 patients were classified according to WHO disease severity. We evaluated the levels of IFN-α, IFN-γ, TNF-α, IL-1R, IL-6, IL-7, IL-10, CCL2, CCL3, CXCL8, CXCL10 and GCSF in the serum and were compared among COVID-19 disease severity groups and stratified by polymorphism alleles. This study revealed a significant increase in IL-2, IL-6 and CCL-2 levels in the dead group. However, the IL-10 levels were higher in the moderate group than in the mild group. Logistic regression analysis revealed that five polymorphisms were associated with an increased risk of severe COVID-19: the *TNF*-α (rs1800610) A allele (OR=1.50; 95% CI: 1.01–2.24); the *IL-6* (rs1800796) C allele (OR=1.64; 95% CI: 1.05–2.57); the *IL-10* (rs1800871) T allele (OR=1.94; 95% CI: 1.24–3.04) and the *IL-10* (rs1800872) A allele (OR=1.87; 95% CI: 1.21–2.89); and the CCL5 (rs3817656) G allele (OR= 1.64; 95% CI: 1.02–2.65)). The *IL-10* (rs1800871 and rs1800872) and *IL-6* (rs1800796 and rs18049563) gene polymorphisms were also associated with COVID-19 severity. Increases in IL-6 and CCL-2 serum levels in carriers of the risk allele rs1049953. In contrast, the IL-10 levels were not associated with any of the SNPs.

## Introduction

Severe acute respiratory syndrome coronavirus 2 (SARS-CoV-2) is the etiologic agent of coronavirus disease 2019 (COVID-19). Some people with this disease can develop acute respiratory distress syndrome (ARDS). Their clinical manifestations have been described in diverse scenarios, ranging from asymptomatic to severe outcomes, and are characterized by hyperinflammation. In COVID-19, an immune response involving early proinflammatory cytokines such as IFN-α, IFN-γ, IL-10, and IL-6 or acute-phase proteins such as C-reactive protein (CRP), among other cytokines such as monocyte chemotactic protein-3 (MCP-1/CCL-2), has been described (1, 2).

In addition, different risk factors involved in outcome severity have also been described, including older age, male sex, obesity, and cardiovascular disease, among others (3). However, these risk factors do not explain all severe cases; genetic host variation might influence the clinical outcomes of patients with COVID-19 (4). Furthermore, previous studies have reported the presence of host genetic variation that might have a high impact on the clinical outcomes of patients with COVID-19 (4). These genetic variations may affect innate and adaptive immune responses (5, 6). Polymorphisms in genes encoding proinflammatory cytokines may play a role in the pathogenesis of severe COVID-19 and could even influence their production.

Recently, single nucleotide polymorphisms (SNPs) in the *TNF-*α, *IL-6, IL-8, IL-10*, *CCL5* and *CXCL6* genes have been found to be associated with COVID-19 severity, suggesting that SNPs could help explain COVID-19 outcomes (7, 8). In this context, TNF-α functions as a mediator of innate immunity, and as an important part of the cellular immune response it can be secreted by different immune cells, such as monocytes, lymphocytes, fibroblasts, etc. (9); however, high concentrations of TNF-α are associated with hepatitis C virus (HCV) infection and may influence COVID-19 severity.

In addition, IL-6 is a cytokine that affects the proinflammatory and anti-inflammatory functions of cellular immunity (10), whereas increased levels of this cytokine have been detected in severe illnesses such as sepsis (11) and ARDS caused by SARS-CoV-2 (12, 13).

IL-8 is a proinflammatory cytokine that consists of two peptides, Ser-IL-8 and Ala-IL-8, and has been associated with pulmonary disease, bladder cancer and rheumatoid arthritis (RA) (14). Elevated serum levels of this cytokine have been observed in patients with severe COVID-19 (15–17).

Meanwhile, IL-10 is a cytokine that plays a role in chronic viral infections and presents intriguing therapeutic challenges (18). This cytokine promotes immune-mediated eradication of viruses, such as cytomegalovirus, lymphocytic choriomeningitis virus, human immunodeficiency virus (HIV) and HCV (19).

Chemokine (C-X-C-motif (CXCs)) ligand 6 (CXCL6) induces angiogenesis via CXCR2 under chronic hypoxia (20) and has been observed to play a role in some diseases, such as prostate cancer and acute myeloid leukemia (21). CXCL6 may influence the severity of COVID-19 by its role in hypoxia (22).

Finally, CCL5, also called RANTES, is a chemokine with a high affinity for CCR5, a chemokine receptor that also binds to CCL3 and CCL4. Low CCL5 expression levels in the upper respiratory tract have been reported in patients with high SARS-CoV-2 viral loads, intensive care unit (ICU) admission, and death (23).

The aim of this work was to evaluate the associations of polymorphisms in the *TNF*-α*, IL-6*, *IL-8, IL-10*, *CCL5* and *CXCL6* genes with COVID-19 outcomes and with the serum levels of IFN-α, IFN-γ, TNF-α, IL-1Ra, IL-6, IL-7, IL-10, CCL2, CCL3, CXCL8, CXCL10 and GCSF.

## Materials and methods

### Setting and participants

From June 2020 to March 2021, nonprobability sampling was performed with unvaccinated patients in a multicenter cross-sectional study from different Mexican governmental health care institutions. This study was conducted in accordance with good clinical practice and the Declaration of Helsinki. Informed consent was obtained from each participant before they entered the study. This study was approved by the ethics committee of the Instituto Nacional de Rehabilitación Luis Guillermo Ibarra Ibarra (INR-LGII: 17/20). The inclusion criteria were as follows: either sex, age ≥18 years with clinical features of COVID-19, and positive qRT–PCR test from a nasopharyngeal swab. The exclusion criteria were pregnancy, incomplete clinical records and individuals related to included participants. The participants were classified according to the WHO clinical progression scale and scored as follows: ambulatory mild disease (score of 1-3), hospitalized moderate disease (score of 4-5), severe disease (score of 6-9) and death (score of 10) (24).

### Selection and genotyping of SNPs

Peripheral blood samples were collected from each participant at the hospital’s COVID-19 triage for DNA and serum isolation. Genomic DNA was isolated using a specialized commercial kit (QIAmp DNA Blood Mini Kit, Qiagen, Hilden, Germany). The quality of the DNA samples was evaluated by the 260/280 nm absorbance ratio, and 1% agarose gels were stained with SYBR® Green (Invitrogen, CA, USA). The DNA concentration was subsequently quantified using a spectrophotometric method and adjusted to 20 ng/μl. In addition, a vacutainer tube with SST II Advance gel was used for serum isolation. The serum samples were separated and stored at −80 °C until further use.

The polymorphisms of *TNF*-α (rs1800610 (+489 G/A), rs1800629 (−308 G/A), and rs3093664), *IL-6* (rs1800796 (−572 G/C) and rs10499563 (−6331 T/C)), *IL-8* (rs2227307), *IL-10* (rs1800872 (−819 C/T) and rs1800071 (−592 C/A)), *CXCL6* (rs4279174), and *CCL5* (rs2107538 and rs3817656) were selected on the basis of their previous scientific evidence of associations with different diseases in any population that included independent genetic studies from 2003–2020. The included polymorphisms all had a minor allele frequency (MAF) ≥5% according to the 1000 Genomes Project or Hap map for the Mexican population (MXL) or the Iberia (IBS) population.

For genotyping, 10 ng/μl of genomic DNA was transferred into OpenArray plates, which previously contained the specific genotyping primers and probes, via the AccuFill system. Real-time PCR amplification was performed according to the supplier’s protocol via the OpenArray Platform through a Quant Studio 12K Flex System (Thermo Fisher Scientific, Waltham, MA, USA), and the results were analyzed using TaqMan Genotyper v1.6 software.

### Soluble molecule evaluation

Serum IFN-α, IFN-γ, TNF-α, IL-1Ra, IL-6, IL-7, IL-10, CCL2, CCL3, CXCL8, CXCL10 and GCSF levels were measured using a Human COVID-19 Cytokine Storm Panel 1 (14 plex) provided by Biolegend (Cat. No. 741088, Lot B377360) following the manufactureŕs instructions.

### Statistical analysis

The normality of the variable distributions was evaluated. The Kruskal Wallis test was used to compare nonparametric continuous variables among the studied groups, and the results are presented as the median and interquartile range (IQR). For the categorical variables, the chi-square test was performed. For all tests, a value of p <0.05 was considered to indicate statistical significance.

In addition, the linkage disequilibrium (LD) among all variants was determined using HaploView software V4.2.

Logistic regression analysis adjusted for age, sex, hypertension status, type 2 diabetes status and obesity status was used to evaluate associations between genetic variants and the outcomes of COVID-19 patients. The final models were evaluated using the Hosmer– Lemeshow goodness-of-fit test. The analysis was performed using the STATA v.16 statistical package (StataCorp Texas, USA).

## Results

### Study population

In the present work, 627 subjects were included in the analysis and their COVID-19 disease severity was stratified by the WHO criteria as follows: 20% mild (n=123), 36% moderate (n=229), 26% severe (n=162) and 18% dead (n=113). Table 1 shows the features and clinical information of the study population. The median age of the total population was 52 years (IQR 43-63), with older subjects in the deceased group having a median age of 63 years. In the entire population, 63% were males, and the principal comorbidities were obesity in 31% (n=197), hypertension in 30% (n=188) and type 2 diabetes mellitus in 31% (n=193).

**Table 1.**
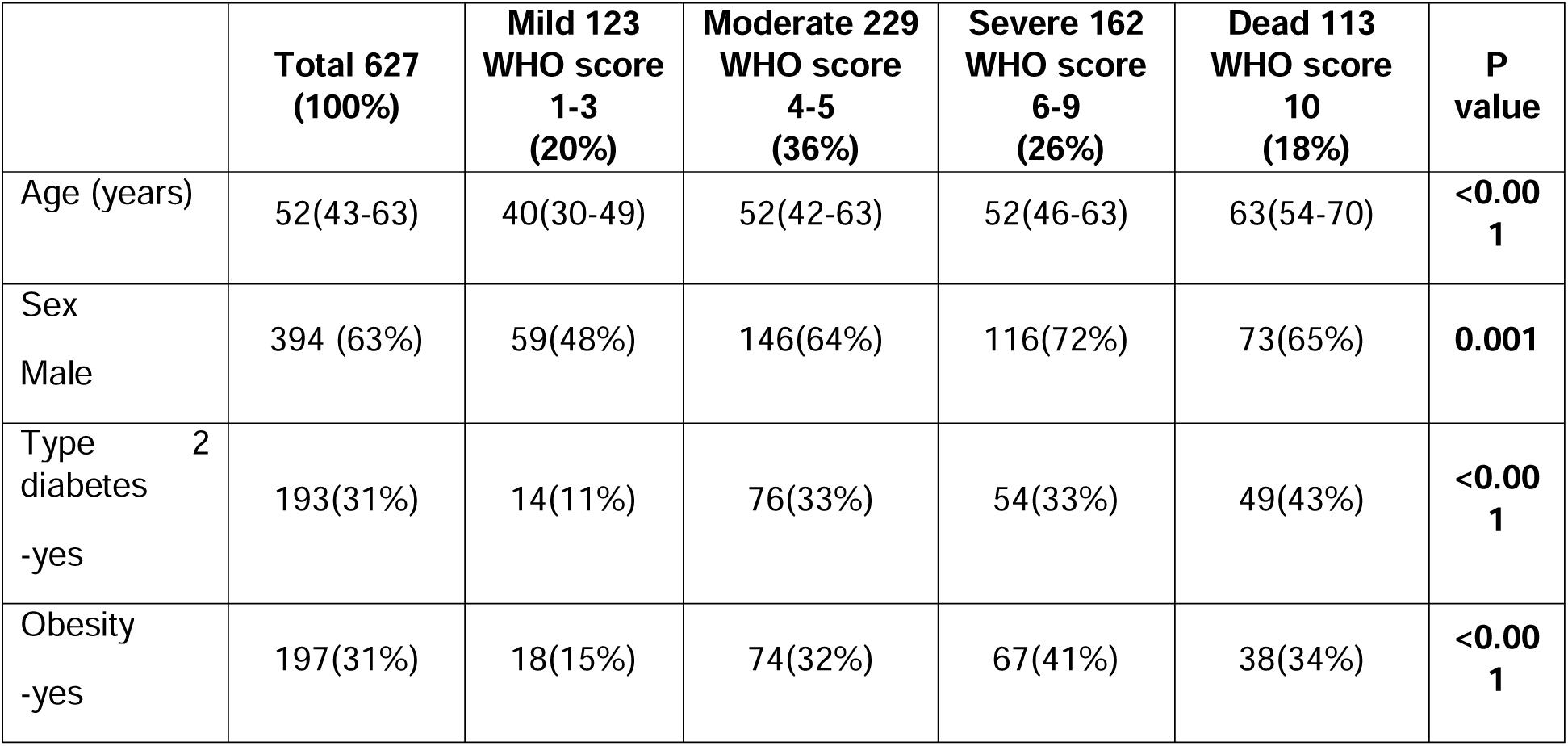
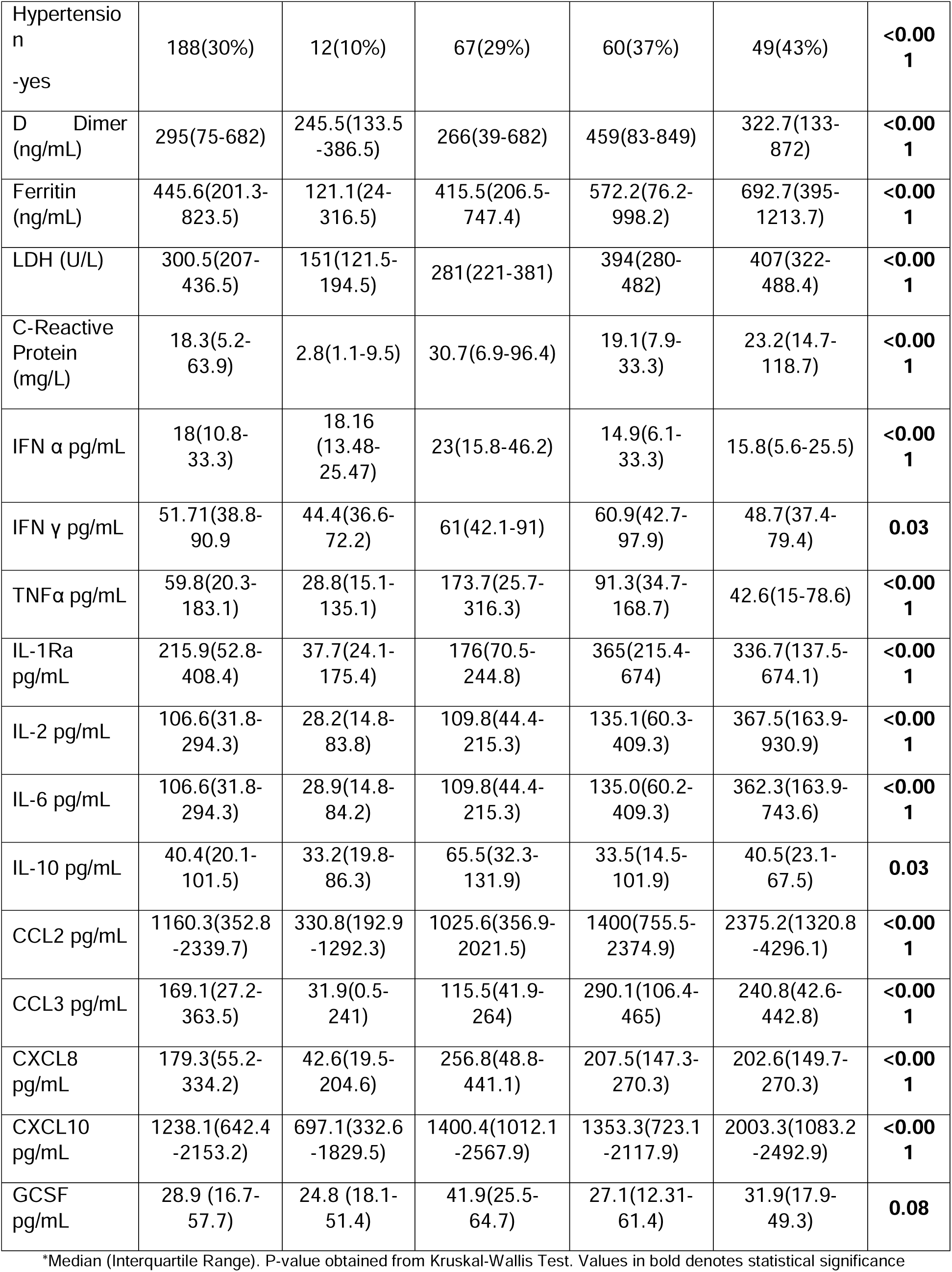
Anthropometric and clinical characteristics of population study.

For clinical biomarkers, we observed statistical significance (*p*<0.001) among the groups, where ferritin, lactate dehydrogenase (LDH) and CRP levels tended to increase with the severity of the disease, with the highest levels occurring in the deceased group (Table 1).

With respect to IL-6 and CCL2, we observed a tendency toward higher levels (pg/mL) as the severity of the disease increased. Similarly, CXCL-10 showed the highest concentration in the deceased group, and IL-1Ra and CCL3 also presented the highest concentrations in the severe and deceased COVID-19 groups. Interestingly, the concentrations of IFN-α, IFN-γ, TNF-α, IL-2, IL-10, CXCL8 and GCSF were the highest in the moderate disease group (Table 1).

### Allelic and genotypic frequencies and linkage disequilibrium

The allelic and genotypic frequencies were calculated and are shown in Table 2. We observed a significant difference in the distribution of the allele frequencies among the study groups for the *IL-6* rs1800796 (−572 G/C) and rs10499563 (−6331 T/C); *IL-10* rs1800871 (−819 T/C) and rs1800872 (−592 C/A); and *CCL5* rs3817656 polymorphisms. The LD was calculated for all SNPs; however, only rs1800871 (−819 T/C) and rs1800872 (−592 C/A) of the *IL-10* gene showed LD, with R^2^ values of 0.99 and D’=0.99, as did rs2107538 and rs3817656 of CCL5 (R^2^ values of 0.79 and D’=0.95).

**Table 2.** Allelic and Genotypes Frequencies in Mild, Moderate, Severe, and Dead patients COVID-19.

|  | **Total 627**<br>**(100%)** | **Mild 123**<br>**(20%)** | **Moderate**<br>**229 (36%)** | **Severe 162**<br>**(26%)** | **Dead**<br>**113 (18%)** | ***P***<br>**value\*\*** |
| --- | --- | --- | --- | --- | --- | --- |
| **TNF-a** |  |  |  |  |  |  |
| rs1800610<br>(+489 G/A) |  |  |  |  |  |  |
| G | 864 (73%) | 175 (76%) | 307 (69%) | 224 (75%) | 158 (73%) | 0.19 |
| A | 326 (27%) | 55 (24%) | 137 (31%) | 76 (25%) | 58 (27%) |  |
| GG | 322(54%) | 70(61%) | 107(48%) | 84(56%) | 61(56%) | 0.37 |
| GA | 222(37%) | 35(30%) | 93(42%) | 56(37%) | 38(35%) |  |
| AA | 52(9%) | 10(9%) | 22(10%) | 10(7%) | 10(9%) |  |
| rs1800629<br>(-308 G/A) |  |  |  |  |  |  |
| G | 1144 (95%) | 209 (92%) | 421 (95%) | 307 (97%) | 207 (96%) | 0.06 |
| A | 60 (5%) | 19 (8%) | 21 (5%) | 11 (3%) | 9 (4%) |  |
| GG | 545(90%) | 95(83%) | 202(91%) | 148(93%) | 100(92%) | 0.05 |
| GA | 56(9%) | 19(17%) | 17(8%) | 11(7%) | 9(8%) |  |
| AA | 2(.33%) | 0(0%) | 2(1%) | 0(0%) | 0(0%) |  |
| rs3093664 |  |  |  |  |  |  |
| A | 1096 (95%) | 205 (95%) | 425 (96%) | 263 (95%) | 203 (95%) | 0.91 |
| G | 56 (5%) | 11 (5%) | 19 (4%) | 15 (5%) | 11 (5%) |  |
| AA | 524 (91%) | 97 (90%) | 204 (92%) | 125 (90%) | 98 (91%) | 0.63 |
| AG | 48 (8%) | 11 (10%) | 17 (8%) | 13 (9%) | 7 (7%) |  |
| GG | 4 (1%) | 0 | 1 (0.5%) | 1 (1%) | 2 (2%) |  |
| IL-6 |  |  |  |  |  |  |
| rs1800796<br>(-572 G/C) |  |  |  |  |  |  |
| G | 725(62%) | 164(68%) | 271(62%) | 173(61%) | 117(55%) | 0.04 |
| C | 445(38%) | 76(32%) | 163(38%) | 111(39%) | 95(45%) |  |
| GG | 231(39%) | 56(47%) | 92(42%) | 55(39%) | 28(26%) | 0.02 |
| GC | 263(45%) | 52(43%) | 87(40%) | 63(44%) | 61(58%) |  |
| CC | 91(16%) | 12(10%) | 38(18%) | 24(17%) | 17(16%) |  |
| rs10499563<br>(-6331 T/C) |  |  |  |  |  |  |
| T | 973(83%) | 196(81%) | 360(83%) | 229(81%) | 188(88%) | 0.15 |
| C | 203(17%) | 46(19%) | 76(17%) | 55(19%) | 26(12%) |  |
| TT | 424(72%) | 84(69%) | 163(75%) | 93(65%) | 84(78%) | 0.007 |
| TC | 125(21%) | 28(23%) | 34(16%) | 43(30%) | 20(19%) |  |
| CC | 39(7%) | 9(7%) | 21(10%) | 6(4%) | 3(3%) |  |
| IL-8 |  |  |  |  |  |  |
| rs2227307 |  |  |  |  |  |  |
| T | 815 (71%) | 151 (70%) | 317 (72%) | 199 (70%) | 148 (70%) | 0.88 |
| G | 311 (28%) | 67 (31%) | 123 (28%) | 85 (30%) | 62 (29%) |  |
| TT | 291(50%) | 51(47%) | 118(54%) | 68(48%) | 54(51%) | 0.74 |
| TG | 234(41%) | 49(45%) | 81(37%) | 63(44%) | 41(39%) |  |
| GG | 52(9%) | 9(8%) | 21(10%) | 11(10%) | 11(10%) |  |
| IL-10 |  |  |  |  |  |  |
| rs1800871<br>(-819 C/T) |  |  |  |  |  |  |
| C | 666(58%) | 131(68%) | 256(59%) | 158(51%) | 121(58%) | 0.002 |
| T | 484(42%) | 63(32%) | 176(41%) | 156(49%) | 89(42%) |  |
| CC | 195(34%) | 48(50%) | 73(34%) | 42(27%) | 32(31%) | 0.001 |
| CT | 276(48%) | 35(36%) | 110(51%) | 74(47%) | 57(54%) |  |
| TT | 104(18%) | 14(14%) | 33(15%) | 41(26%) | 16(15%) |  |
| rs1800872<br>(-592 C/A) |  |  |  |  |  |  |
| C | 677(57%) | 141(67%) | 253(58%) | 162(51%) | 121(56%) | 0.005 |
| A | 503(43%) | 71(33%) | 181(42%) | 156(49%) | 95(44%) |  |
| CC | 198(34%) | 48(49%) | 73(34%) | 42(27%) | 32(31%) | 0.001 |
| CA | 276(48%) | 35(36%) | 110(51%) | 74(47%) | 57(54%) |  |
| AA | 104(18%) | 14(14%) | 33(15%) | 41(26%) | 16(15%) |  |
| CXCL6 |  |  |  |  |  |  |
| rs4279174 |  |  |  |  |  |  |
| G | 914 (78%) | 179 (75%) | 342 (80%) | 229 (76%) | 164 (76%) | 0.47 |
| C | 260 (22%) | 59 (25%) | 86 (20%) | 63 (22%) | 52 (24%) |  |
| GG | 364(62%) | 67(56%) | 143(67%) | 90(62%) | 64(59%) | 0.30 |
| GC | 190(32%) | 47(39%) | 56(26%) | 49(34%) | 38(35%) |  |
| CC | 35(6%) | 6(5%) | 15(7%) | 7(5%) | 35(6%) |  |
| CCL5 |  |  |  |  |  |  |
| rs2107538 |  |  |  |  |  |  |
| C | 784 (68%) | 148 (73%) | 289 (66%) | 193 (66%) | 154 (71%) | 0.25 |
| T | 364 (32%) | 56 (27%) | 147 (34%) | 99 (34%) | 62 (29%) |  |
| CC | 270(47%) | 50(49%) | 94(43%) | 68(47%) | 58(53%) | 0.10 |
| CT | 246(43%) | 48(47%) | 101(46%) | 57(39%) | 40(37%) |  |
| TT | 59(10%) | 4(4%) | 23(11%) | 21(14%) | 11(10%) |  |
| rs3817656 |  |  |  |  |  |  |
| A | 817 (72%) | 179 (80%) | 282 (70%) | 194 (68%) | 162 (76%) | **0.009** |
| G | 311 (28%) | 45 (20%) | 124 (30%) | 90 (32%) | 52 (24%) |  |
| AA | 306(54%) | 68(60%) | 105(52%) | 72(51%) | 61(56%) | **0.004** |
| AG | 208(37%) | 44(39%) | 72(36%) | 50(35%) | 42(39%) |  |
| AA | 52(9%) | 1(1%) | 26(13%) | 20(14%) | 5(5%) |  |

\*\* Chi-square test, values in bold denotes statistical significance

### Logistic regression analysis

For the *TNF-*α gene, we observed a significant association of the rs1800610 risk allele with moderate disease (OR=1.5, 95% CI= 1.01-2.24, *p*=0.04) and the rs1800629 minor allele with moderate disease (OR=0.45, 95% CI95% CI= 0.22-0.94, *p*=0.03) and severe outcomes (OR=0.34, 95% CI95% CI=0.14-0.83, *p*=0.02) (Table 3).

**Table 3.** Association of the Polymorphisms of *TNF*α (rs1800610, rs1800629, rs3093664), *IL-6* (rs1800796, rs10499563), *IL-8* (rs2227307), *IL-10* (rs1800872 and rs1800071), *CXCL6* (rs4279174) and *CCL5* (rs2107538, rs3817656) genes and COVID-19 outcomes.

| Polymorphisms | Moderate |  |  | Severe |  |  | Dead |  |  |
| --- | --- | --- | --- | --- | --- | --- | --- | --- | --- |
|  | OR* | 95% CI | P value | OR* | 95% CI | P value | OR* | 95% CI | P value |
| **TNF-α** |  |  |  |  |  |  |  |  |  |
| rs1800610<br>(+489 G/A) |  |  |  |  |  |  |  |  |  |
| G | Reference |  |  | Reference |  |  | Reference |  |  |
| A | **1.50** | **1.01-2.24** | **0.04** | 1.11 | 0.69-1.79 | 0.65 | 1.30 | 0.74-2.28 | 0.35 |
| GG<sup>c</sup> | Reference |  |  | Reference |  |  | Reference |  |  |
| GA<sup>c</sup> | 1.72 | 1.01-2.96 | 0.05 | 1.16 | 0.62-2.17 | 0.65 | 1.11 | 0.52-2.37 | 0.79 |
| AA<sup>c</sup> | 1.68 | .067-4.08 | 0.25 | 1.13 | 0.37-3.43 | 0.83 | 1.70 | 0.51-5.67 | 0.39 |
| GA+AA<sup>d</sup> | **1.72** | **1.04-2.84** | **0.04** | 1.15 | 0.64-2.08 | 0.64 | 1.22 | 0.61-2.47 | 0.57 |
| AA<sup>r</sup> | 1.36 | 0.58-3.21 | 0.48 | 1.08 | 0.36-3.20 | 0.89 | 1.64 | 0.51-5.31 | 0.41 |
| rs1800629<br>(-308 G/A) |  |  |  |  |  |  |  |  |  |
| G | Reference |  |  | Reference |  |  | Reference |  |  |
| A | **0.45** | **0.22-0.94** | **0.03** | **0.34** | **0.14-0.83** | **0.02** | 0.52 | 0.20-1.48 | 0.22 |
| rs3093664 |  |  |  |  |  |  |  |  |  |
| A | Reference |  |  | Reference |  |  | Reference |  |  |
| G | 0.87 | 0.37-2.06 | 0.76 | 0.99 | 0.38-2.59 | 0.99 | 1.06 | 0.35-3.23 | 0.91 |
| **IL-6** |  |  |  |  |  |  |  |  |  |
| rs1800796<br>(-572 G/C) |  |  |  |  |  |  |  |  |  |
| G | Reference |  |  | Reference |  |  | Reference |  |  |
| C | 1.35 | 0.92-1.97 | 0.12 | **1.64** | **1.05-2.57** | **0.03** | 1.34 | 0.77-2.34 | 0.3 |
| GG<sup>c</sup> | Reference |  |  | Reference |  |  | Reference |  |  |
| GC<sup>c</sup> | 1.03 | 0.60-1.79 | 0.89 | 1.34 | 0.68-2.67 | 0.38 | 1.76 | 0.76-4.08 | 0.18 |
| CC<sup>c</sup> | 1.87 | 0.83-4.21 | 0.12 | **2.83** | **1.11-7.26** | **0.03** | 1.53 | 0.45-5.18 | 0.48 |
| GC+CC<sup>d</sup> | 1.20 | 0.72-2.00 | 0.48 | 1.64 | 0.87-3.07 | 0.12 | 1.71 | 0.77-3.77 | 0.18 |
| CC<sup>r</sup> | 1.84 | 0.86-3.95 | 0.12 | **2.44** | **1.01-5.87** | **0.04** | 1.13 | 0.36-3.53 | 0.83 |
| rs10499563<br>(-6331 T/C) |  |  |  |  |  |  |  |  |  |
| T | Reference |  |  | Reference |  |  | Reference |  |  |
| C | 1.04 | 0.65-1.66 | 0.84 | 1.25 | 0.72-2.18 | 0.41 | 0.74 | 0.34-1.62 | 0.46 |
| TT<sup>c</sup> | Reference |  |  | Reference |  |  | Reference |  |  |
| TC<sup>c</sup> | 0.69 | 0.36-1.33 | 0.27 | 1.87 | 0.91-3.86 | 0.09 | 0.78 | 0.28-2.13 | 0.63 |
| CC<sup>c</sup> | 1.58 | 0.63-3.93 | 0.38 | 0.73 | 0.19-2.79 | 0.65 | 0.65 | 0.11-3.64 | 0.62 |
| TC+CC<sup>d</sup> | 0.91 | 0.52-1.59 | 0.74 | 1.56 | 0.80-3.04 | 0.18 | 0.74 | 0.30-1.86 | 0.53 |
| CC<sup>r</sup> | 1.69 | 0.68-4.18 | 0.25 | 0.62 | 0.17-2.31 | 0.48 | 0.68 | 0.12-3.78 | 0.66 |
| **IL-8** |  |  |  |  |  |  |  |  |  |
| rs2227307 |  |  |  |  |  |  |  |  |  |
| T | Reference |  |  | Reference |  |  | Reference |  |  |
| G | 0.91 | 0.61-1.34 | 0.63 | 0.82 | 0.52-1.31 | 0.42 | 0.78 | 0.45-1.34 | 0.37 |
| TT<sup>c</sup> | Reference |  |  | Reference |  |  | Reference |  |  |
| TG<sup>c</sup> | 0.71 | 0.41-1.22 | 0.22 | 0.86 | 0.46-1.60 | 0.63 | 0.63 | 0.29-1.34 | 0.24 |
| GG<sup>c</sup> | 1.11 | 0.44-2.83 | 0.81 | 0.61 | 0.19-1.96 | 0.41 | 0.84 | 0.24-2.94 | 0.79 |
| TG+GG<sup>d</sup> | 0.77 | 0.47-1.29 | 0.32 | 0.82 | 0.45-1.49 | 0.51 | 0.67 | 0.33-1.36 | 0.27 |
| GG<sup>r</sup> | 1.30 | 0.53-3.19 | 0.57 | 0.66 | 0.21-2.03 | 0.47 | 1.02 | 0.31-3.41 | 0.97 |
| **IL-10** |  |  |  |  |  |  |  |  |  |
| rs1800871<br>(-819 C/T) |  |  |  |  |  |  |  |  |  |
| C | Reference |  |  | Reference |  |  | Reference |  |  |
| T | 1.24 | 0.83-1.85 | 0.28 | **1.94** | **1.24-3.04** | **0.004** | 1.49 | 0.84-2.63 | 0.16 |
| CC<sup>c</sup> | Reference |  |  | Reference |  |  | Reference |  |  |
| CT<sup>c</sup> | 1.78 | 0.98-3.24 | 0.06 | 2.01 | 0.98-4.09 | 0.05 | **3.7** | **1.47-9.19** | **0.005** |
| TT<sup>c</sup> | 1.24 | 0.56-2.78 | 0.06 | **3.00** | **1.28-7.00** | **0.01** | 1.41 | 0.42-4.72 | 0.57 |
| CT+TT<sup>d</sup> | 1.62 | 0.94-2.82 | 0.08 | **2.33** | **1.21-4.44** | **0.01** | **2.85** | **1.22-6.61** | **0.02** |
| TT<sup>r</sup> | 0.93 | 0.43-1.94 | 0.83 | 2.09 | 0.97-4.48 | 0.06 | 0.71 | 0.23-2.08 | 0.53 |
| rs1800872<br>(-592 C/A) |  |  |  |  |  |  |  |  |  |
| C | Reference |  |  | Reference |  |  | Reference |  |  |
| A | 1.23 | 0.84-1.82 | 0.27 | **1.87** | **1.21-2.89** | **0.004** | **1.73** | **1.01-2.9** | **0.04** |
| CC<sup>c</sup> | Reference |  |  | Reference |  |  | Reference |  |  |
| CA<sup>c</sup> | 1.64 | 0.92-2.93 | 0.09 | 1.83 | 0.92-3.64 | 0.08 | **3.17** | **1.32-7.58** | **0.009** |
| AA<sup>c</sup> | 1.34 | 0.62-2.91 | 0.45 | **2.89** | **1.26-6.60** | **0.01** | 2.31 | 0.74-7.15 | 0.14 |
| CA+AA<sup>d</sup> | 1.56 | 0.91-2.66 | 0.11 | **2.16** | **1.15-4.03** | **0.02** | **2.91** | **1.28-6.60** | **0.01** |
| AA<sup>r</sup> | 1.03 | 0.51-2.11 | 0.92 | **2.11** | **1.01-4.44** | **0.04** | 1.19 | 0.44-3.22 | 0.73 |
| Haplotypes |  |  |  |  |  |  |  |  |  |
| CC | Reference |  |  | Reference |  |  | Reference |  |  |
| TA | 1.26 | 0.84-1.89 | 0.25 | **1.97** | **1.25-3.08** | **0.003** | 1.54 | 0.87-2.7 | 0.14 |
| **CXCL6** |  |  |  |  |  |  |  |  |  |
| rs4279174 |  |  |  |  |  |  |  |  |  |
| G | Reference |  |  | Reference |  |  | Reference |  |  |
| C | 0.78 | 0.52-1.19 | 0.26 | 0.72 | 0.44-1.15 | 0.17 | 0.82 | 0.46-1.45 | 0.50 |
| GG<sup>c</sup> | Reference |  |  | Reference |  |  | Reference |  |  |
| GC<sup>c</sup> | 0.58 | 0.33-0.99 | 0.05 | 0.65 | 0.35-1.19 | 0.17 | 0.65 | 0.31-1.33 | 0.24 |
| CC<sup>c</sup> | 1.28 | 0.43-3.79 | 0.66 | 0.66 | 0.17-2.60 | 0.55 | 1.12 | 0.25-4.98 | 0.88 |
| GC+CC<sup>d</sup> | 0.65 | 0.39-1.08 | 0.10 | 0.65 | 0.36-1.17 | 0.15 | 0.70 | 0.35-1.40 | 0.31 |
| CC<sup>r</sup> | 1.55 | 0.53-4.53 | 0.42 | 0.77 | 0.20-2.97 | 0.71 | 1.30 | 0.30-5.66 | 0.73 |
| **CCL5** |  |  |  |  |  |  |  |  |  |
| rs2107538 |  |  |  |  |  |  |  |  |  |
| C | Reference |  |  | Reference |  |  | Reference |  |  |
| T | 1.31 | 0.87-1.95 | 0.19 | 1.22 | 0.76-1.94 | 0.39 | 1.36 | 0.78-2.36 | 0.27 |
| CC<sup>c</sup> | Reference |  |  | Reference |  |  | Reference |  |  |
| CT<sup>c</sup> | 0.94 | 0.54-1.62 | 0.83 | 0.73 | 0.39-1.39 | 0.35 | 0.74 | 0.34-1.60 | 0.45 |
| TT<sup>c</sup> | **3.39** | **1.04-10.9** | **0.04** | 3.27 | 0.92-11.58 | 0.06 | **4.83** | **1.14-20.39** | **0.03** |
| CT+TT<sup>d</sup> | 1.13 | 0.67-1.90 | 0.66 | 0.95 | 0.52-1.73 | 0.86 | 1.01 | 0.49-2.07 | 0.98 |
| TT<sup>r</sup> | **3.50** | **1.11-10.95** | **0.03** | **3.75** | **1.10-12.84** | **0.04** | **5.48** | **1.35-22.37** | **0.02** |
| rs3817656 |  |  |  |  |  |  |  |  |  |
| A | Reference |  |  | Reference |  |  | Reference |  |  |
| G | **1.66** | **1.08-2.54** | **0.02** | **1.64** | **1.02-2.65** | **0.04** | 1.50 | 0.84-2.67 | 0.16 |
| AA<sup>c</sup> | Reference |  |  | Reference |  |  | Reference |  |  |
| AG<sup>c</sup> | 0.87 | 0.51-1.50 | 0.62 | 0.82 | 0.43-1.54 | 0.55 | 1.17 | 0.57-2.43 | 0.66 |
| GG<sup>c</sup> | **14.76** | **1.91-113.7** | **0.01** | **15.63** | **1.92-127.0** | **0.01** | 11.29 | 1.00-127.2 | 0.50 |
| AG+GG<sup>d</sup> | 1.22 | 0.73-2.04 | 0.45 | 1.21 | 0.67-2.18 | 0.52 | 1.36 | 0.67-2.75 | 0.39 |
| GG<sup>r</sup> | **15.52** | **2.04-118.3** | **0.008** | **16.74** | **2.09-134.3** | **0.008** | 10.57 | 0.96-116.8 | 0.05 |
| Haplotypes |  |  |  |  |  |  |  |  |  |
| CA | Reference |  |  | Reference |  |  | Reference |  |  |
| TG | 1.39 | 0.87-2.20 | 0.17 | 1.31 | 0.76-2.24 | 0.33 | 1.35 | 0.68-2.67 | 0.38 |

For *IL-6,* we found a significant positive association for the rs1800796 (−572 G/C) C allele with severe outcomes, with an OR of 1.64 (95% CI=1.05–2.57, *p*=0.03). Additionally, for the CC genotype of the codominant model, the OR was 2.83 (95% CI=1.11–7.26; *p*=0.03) for severe outcomes. In addition, for the CC of the recessive model, we found an OR of 2.44 (95% CI=1.01-5.87, *p*=0.04) for severe outcomes (Table 3). For rs1049953 (−6331T/C), we did not observe a significant association with any outcome.

Interestingly, for the *IL-10* gene rs1800871 (−819 C/T) polymorphism, we observed a significant positive association between the risk allele and severe outcomes, with an OR of 1.94 (95% CI=1.24-3.04, p=0.004). The TT genotype of the codominant model had an OR=3.00 (95% CI=1.28-7.00, *p*=0.01), and the CT+TT genotypes of the dominant model had an OR=2.33 (95% CI 1.21-4.44, *p*=0.01) for severe outcomes. In addition, for the deceased outcome, we observed a significant association with CT in the codominant model (OR=3.7 95% CI=1.47-9.19, *p*=0.005) and CT+TT in the dominant model (OR=2.85 95% CI=1.22-6.61, *p*=0.02). For rs1800872 (−592 C/A), the A allele was associated with severe and deceased outcomes, with ORs of 1.87 (95% CI=1.21-2.89, *p*=0.004) and 1.73 (95% CI=1.01-2.9, *p*=0.04), respectively. The AA genotype of the codominant model had an OR of 2.89 (95% CI=1.26-6.60, *p*=0.01), the CA+AA genotype of the dominant model had an OR=2.16 (95% CI=1.15-4.03, *p*=0.02), and the AA genotype of the recessive model had an OR=2.11 (95% CI =1.01-4.44, *p*=0.04), all of which were associated with severe outcomes. In addition, we observed a significant association between the CA genotype of the codominant model and the CA+AA genotypes of the dominant model with a deceased outcome, with ORs of 3.17 (95% CI=1.32-7.58, *p*=0.009) and 2.91 (95% CI=1.28-6.60, *p*=0.01), respectively.

Because rs1800871 (−819 C/T) and rs1800872 (−592 C/A) were in LD, we performed logistic regression with the haplotypes and found a significant association between the CT haplotype and severe outcomes, with an OR=1.97 (95% CI=1.25-3.08, *p*=0.003) (Table 3). For the *CCL5* gene, the variant rs2107538 in the codominant model (AA genotype) was significantly associated with moderate (OR= 3.39 95% CI= 1.04-10.9, *p=*0.04) and deceased (OR= 4.83 95% CI= 1.14-20.39, *p=*0.03) outcomes. The recessive model (TT genotype) tended to increase the associations among the COVID-19 outcomes (Table 3). Variant rs3817656, the G allele, was significantly associated with moderate (OR= 1.66 95% CI= 1.08-2.54, *p=*0.02) and severe (OR= 1.64 95% CI= 1.02-2.65, *p=*0.04) COVID-19 outcomes (Table 3).

### Cytokine levels and SNPs

We performed a subanalysis to determine the relationships of the concentrations of the cytokines IFN-α, IFN-γ, TNF-α, IL-1Ra, IL-2, IL-6, IL-7, IL-10, CCL-2, CCL3, GCSF, CXCL8, and CXCL10 with the SNPs. In this analysis, the carriers of the risk allele of the *TNF-*α rs1800629 polymorphism presented lower levels of IL-1Ra, IL-6, CCL-2, and CXCL10 than did the carriers of the major allele (Table 4).

**Table 4.** Cytokines concentrations between ancestral and minnor alleles of the polymorphisms of *TNF*α (rs1800610, rs1800629, rs3093664), *IL-6* (rs1800796, rs10499563), *IL-8* (rs2227307), *IL-10* (rs1800872 and rs1800071), *CXCL6* (rs4279174), and *CCL5* (rs2107538, rs3817656) genes.

|  | Total | Ancestral allele | Variant allele | P value |
| --- | --- | --- | --- | --- |
| ***TNF α*** |  |  |  |  |
| **rs1800610 (+489 G/A)** | **Total** | **G** | **A** | **P value** |
| TNFα pg/mL | 59.2(20.3-183.1) | 54.5(601.9-2141.2) | 79.9(21.8-256.6) | 0.13 |
| IFN α pg/mL | 17.8(10.8-32.9) | 17.6(10.6-31.4) | 18.9(11.8-45.4) | 0.09 |
| IFN γ pg/mL | 51.6(38.5-89.9) | 51.2(37.4-88.9) | 54.2(41.5-93.1) | 0.27 |
| IL-1Ra pg/mL | 215.3(51.3-405.8) | 204.3(45.6-407.2) | 237.6(79.6-380.9) | 0.37 |
| IL-2 pg/mL | 26.7(16.7-78.7) | 26.1(16.2-70.9) | 27.3(18.4-96.3) | 0.23 |
| IL-6 pg/mL | 108.3(31.8-298.5) | 106.5(30.6-262.2) | 126.9(43.4-357.4) | 0.19 |
| IL-7 pg/mL | 125(73.7-404.8) | 123.2(73.7-356.6) | 133(70.6-530.4) | 0.34 |
| IL-10 pg/mL | 39.9(20.3-101.5) | 159.4(23.2-347.9) | 196.2(31.4-375.8) | 0.05 |
| CCL2 pg/mL | 1160.3(356.9-2297.7) | 1115.9(338.2-2255.7) | 1280.1(436.3-2669.6) | 0.22 |
| CCL3 pg/mL | 169.9(25.5-360.7) | 159.4(23.2-347.9) | 196.2(31.3-375.8) | 0.52 |
| CXCL8 pg/mL | 179.3(54.2-334.2) | 175.1(52.3-309.8) | 204.6(73.8-382.2) | 0.19 |
| CXCL10 pg/mL | 1299.9(651.6-2206.8) | 1230.6(601.9-2141.2) | 1503.9(686.6-2471.1) | 0.06 |
| GCSF pg/mL | 29.6(17.3-54.8) | 28.4(17.6-52.5) | 37.1(16.4-65.3) | 0.17 |
| **rs1800629 (-308 G/A)** | **Total** | **G** | **A** | **P value** |
| TNFα pg/mL | 62.9(20.5-190.5) | 63.7(20.3-191.6) | 43.9(23.7-137.3) | 0.83 |
| IFN α pg/mL | 17.9(10.8-33.7) | 17.9(10.6-33.8) | 17.1(12.4-23.3) | 0.98 |
| IFN γ pg/mL | 51.7(38.8-91.3) | 52.9(39.0-91.3) | 44.9(35.4-89.9) | 0.23 |
| IL-1Ra pg/mL | 217.2(61.3-408.4) | 224.7(64.0-424.1) | 61.0(24.9-220.2) | **0.003** |
| IL-2 pg/mL | 27.0(16.7-79.3) | 27.2(16.9-79.7) | 18.8(13.9-34.7) | 0.11 |
| IL-6 pg/mL | 109.9(33.3-302.7) | 114.3(35.4-308.7) | 43.8(22.9-143.5) | **0.01** |
| IL-7 pg/mL | 127.8(73.9-428.2) | 129.0(75.2-433.9) | 99.3(52.7-164.9) | 0.11 |
| IL-10 pg/mL | 40.4(20.1-101.5) | 41.1(20.3-101.8) | 23.5(18.2-49.9) | 0.05 |
| CCL2 pg/mL | 1171.6(376.9-2361.5) | 1191.7(378.2-2374.9) | 494.9(195.3-1674.5) | **0.02** |
| CCL3 pg/mL | 169.9(27.2-368.3) | 177.8(30.8-368.3) | 85.6(0.5-441.9) | 0.53 |
| CXCL8 pg/mL | 185.5(57.4-338.2) | 188.5(60.3-341.1) | 153.6(24.8-244.4) | 0.18 |
| CXCL10 pg/mL | 1299.9(651.6-2153.2) | 1317.4(660.9-2230.1) | 741.8(333.3-1400.5) | **0.002** |
| GCSF pg/mL | 28.9(16.7-55.0) | 29.6(16.7-57.7) | 24.8(17.6-41.6) | 0.33 |
| **rs3093664** | **Total** | **A** | **G** | **P value** |
| TNFα pg/mL | 59.2(20.3-183.1) | 54.5(19.5-173.7) | 79.9(21.8-256.6) | 0.05 |
| IFN α pg/mL | 17.6(10.6-32.3) | 17.1(10.45-32.6) | 22.5(13.5-31.7) | 0.27 |
| IFN γ pg/mL | 51.2(37.9-88.9) | 51.1(37.6-89.4) | 55.3(41.8-88.9) | 0.51 |
| IL-1Ra pg/mL | 215.3(51.3-405.8) | 204.3(45.5-407.1) | 237.6(79.6-380.9) | 0.84 |
| IL-2 pg/mL | 25.8(16.5-75.5) | 25.7(16.4-73.6) | 34.7(18.5-102.1) | 0.31 |
| IL-6 pg/mL | 111.2(30.6-286.1) | 112.4(30.6-286.2) | 96.3(52.3-197.6) | 0.84 |
| IL-7 pg/mL | 117(70.6-401.9) | 116.4(69.5-397.1) | 181.7(93.1-501.8) | 0.13 |
| IL-10 pg/mL | 39.9(20.3-101.5) | 37.5(20.2-93.5) | 51.2(23.8-131.9) | 0.15 |
| CCL2 pg/mL | 1160.3(352.8-2339.6) | 1153.8(356.9-2350.5) | 1714.5(292.3-2242.6) | 0.97 |
| CCL3 pg/mL | 169.9(25.5-360.7) | 159.3(23.1-347.9) | 196.2(31.3-375.8) | 0.32 |
| CXCL8 pg/mL | 179.3(54.1-334.2) | 175.1(52.3-309.8) | 204.6(73.8-382.1) | 0.41 |
| CXCL10 pg/mL | 1299.9(651.6-2206.7) | 1230.6(601.8-2141.2) | 1503.9(686.6-2471.1) | 0.94 |
| GCSF pg/mL | 28.9(16.5-53.3) | 28.4(10.5-53.8) | 35.2(19.7-50.2) | 0.65 |
| **IL-6** |  |  |  |  |
| **rs1800796 (-572 G/C)** | **Total** | **G** | **C** | **P value** |
| TNFα pg/mL | 54.5(18.6-173.7) | 52.2(20.3-173.7) | 57.4(16.2-173.7) | 0.51 |
| IFN α pg/mL | 17.6(10.7-32.3) | 18.2(11.4-32.9) | 15.9(9.4-30.9) | 0.08 |
| IFN $\gamma$ pg/mL | 51.1(37.8-87.4) | 51.7(37.8-88.9) | 48.7(37.4-84.9) | 0.57 |
| IL-1Ra pg/mL | 214.5(44.8-405.8) | 195.9(44.1-367.3) | 228.9(64.1-458.7) | 0.13 |
| IL-2 pg/mL | 25.6(16.2-73.6) | 25.5(15.9-75.5) | 25.8(16.9-72.8) | 0.79 |
| IL-6 pg/mL | 108.3(29.7-286.1) | 106.4(30.62-286.02) | 119(28.9-286.2) | 0.89 |
| IL-7 pg/mL | 116.2(69.5-395.8) | 125(73.7-404.8) | 11.3(62.5-352.8) | 0.21 |
| IL-10 pg/mL | 37.5(20.1-98.8) | 40.6(20.5-101.9) | 34.8(19.5-94.1) | 0.18 |
| CCL2 pg/mL | 1157.1(342.9-2283-.9) | 1078.9(334.4-2297.7) | 1239.4(361.1-2242.6) | 0.68 |
| CCL3 pg/mL | 166.9(24.9-368.3) | 155.8(24.9-348.9) | 186.7(24.3-386.8) | 0.63 |
| CXCL8 pg/mL | 175.3(50.8-312.5) | 178.9(51.3-324.8) | 173.2(49.5-309.7) | 0.81 |
| CXCL10 pg/mL | 1230.6(629.9-2151.1) | 1237.3(665.9-2151.1) | 1223.8(529.3-2131.3) | 0.54 |
| GCSF pg/mL | 28.7(16.5-54.5) | 29.6(18.2-54.5) | 26.3(15.4-53.1) | 0.38 |
| **rs10499563 (-6331 T/C)** | **Total** | **T** | **C** | **P value** |
| TNFα pg/mL | 57.4(19.5-173.7) | 53.5(18.1-171.8) | 78.8(23.5-209.5) | 0.14 |
| IFN $\alpha$ pg/mL | 17.9(10.8-33.7) | 17.5(10.1-31.4) | 20.6(11.4-50.4) | **0.04** |
| IFN $\gamma$ pg/mL | 51.2(37.8-88.8) | 49.2(37.6-85.8) | 60.6(40.8-102.1) | 0.07 |
| IL-1Ra pg/mL | 214.9(49.6-405.8) | 200.8(46.2-386.6) | 264.4(55.4-488.8) | 0.12 |
| IL-2 pg/mL | 25.7(16.5-73.6) | 25.5(16.1-73.2) | 33.2(18.4-79.7) | 0.09 |
| IL-6 pg/mL | 109.9(30.6-302.7) | 101.3(29.5-262.2) | 184.2(44.8-372.7) | **0.03** |
| IL-7 pg/mL | 116.5(70.6-398.4) | 115.6(68.4-378.2) | 163.9(83.3-487.4) | 0.05 |
| IL-10 pg/mL | 38.9(20.2-100.2) | 36.9(20.1-95.5) | 52.5(24.3-103.9) | 0.07 |
| CCL2 pg/mL | 1160.3(342.9-2361.4) | 1092.2(338.2-2242.6) | 1665.4(436.3-2736.1) | **0.04** |
| CCL3 pg/mL | 169.2(24.9-360.7) | 154.8(23.1-356.7) | 240.8(77.3-375.8) | 0.09 |
| CXCL8 pg/mL | 175.3(52.3-312.5) | 173.9(49.2-310.7) | 204.9(78.9-390.8) | 0.10 |
| CXCL10 pg/mL | 1238.2(629.9-2206.7) | 1230.6(601.9-2179.9) | 1424.5(688.3-2652.5) | 0.24 |
| GCSF pg/mL | 28.9(16.9-54.5) | 27.9(16.6-53.2) | 33.45(19.5-60.6) | 0.14 |
| **IL-8** |  |  |  |  |

| **rs2227307** | **Total** | **T** | **G** | **P**<br>**value** |
| --- | --- | --- | --- | --- |
| TNFα pg/mL | 58.1(19.5-175.8) | 51.4(18.3-163.9) | 88.9(22.7-226.4) | 0.05 |
| IFN α pg/mL | 17.6(10.6-32.9) | 16.7(10.3-30.9) | 18.3(10.8-43.1) | 0.20 |
| IFN γ pg/mL | 51.1(37.9-88.9) | 48.9(38.3-84.2) | 58.4(35.6-96.0) | 0.35 |
| IL-1Ra pg/mL | 215.4(51.1-408.4) | 215.1(40.9-408.4) | 215.9(85.4-414.9) | 0.46 |
| IL-2 pg/mL | 26.1(16.7-75.5) | 25.6(16.1-70.8) | 30.9(18.6-80.2) | 0.07 |
| IL-6 pg/mL | 111.2(31.8-302.7) | 108.4(29.9-304.4) | 120.1(44.2-290.2) | 0.62 |
| IL-7 pg/mL | 123.2(73.7-401.9) | 116.0(66.4-371.2) | 135.1(80.8-480.3) | 0.09 |
| IL-10 pg/mL | 38.1(20.1-100.2) | 36.7(19.9-92.8) | 44.8(22.1-105.1) | 0.17 |
| CCL2 pg/mL | 1172.3(356.1-2374.9) | 1126.4(344.2-2357.3) | 1320.8(426.5-2644.5) | 0.21 |
| CCL3 pg/mL | 162.4(24.9-368.3) | 153.8(17.9-352.8) | 214.3(34.6-401.1) | 0.15 |
| CXCL8 pg/mL | 177.0(55.2-324.7) | 173.9(49.8-311.9) | 189(78.9-336.2) | 0.22 |
| CXCL10<br>pg/mL | 1282.9(657.2-2151.1) | 1187.6(605.3-2122.9) | 1430.2(700.4-2463.8) | 0.09 |
| GCSF pg/mL | 28.8(16.5-54.5) | 29.6(16.5-54.6) | 28.1(16.6-54.5) | 0.59 |
| **IL-10** |  |  |  |  |
| **rs1800871**<br>**(-819 C/T)** | **Total** | **C** | **T** | **P**<br>**value** |
| TNFα pg/mL | 66.1(20.7-191.7) | 66.2(19.5-205.1) | 66(21.8-173.7) | 0.70 |
| IFN α pg/mL | 17.9(10.8-34.3) | 17.7(10.9-36.8) | 18.1(9.7-33.3) | 0.39 |
| IFN γ pg/mL | 53.5(39-92.4) | 51.6(38.5-95.8) | 54.4(40.2-85.8) | 0.96 |
| IL-1Ra pg/mL | 226.6(69.2-433.2) | 215.7(53.3-413.2) | 253.3(101.7-437.2) | 0.07 |
| IL-2 pg/mL | 27.4(16.9-81.8) | 27.8(16.6-87.5) | 27.3(17.5-79.3) | 0.99 |
| IL-6 pg/mL | 126.2(36.3-316.8) | 114.3(30.6-332.6) | 126.9(4.8-304.4) | 0.39 |
| IL-7 pg/mL | 130.2(76.4-459.6) | 132(76.1-488) | 128.5(76.4-398.4) | 0.64 |
| IL-10 pg/mL | 40.6(20.3-95.3) | 40.6(19.9-109.3) | 39.04(20.3-95.3) | 0.35 |
| CCL2 pg/mL | 1258.3(436.3-2506.2) | 1172.9(342.9-2506.2) | 1324.8(525.9-2644.4) | 0.14 |
| CCL3 pg/mL | 186.7(32.4-375.8) | 169.1(20.5-362.1) | 217.9(51.4-387.2) | 0.13 |
| CXCL8 pg/mL | 193.6(66.7-343.1) | 189.9(57.4-363.9) | 197.2(78.9-311.5) | 0.77 |
| CXCL10<br>pg/mL | 1322.5(667.1-2206.8) | 1360.9(667.1-2213.7) | 1273.8(666.3-2153.2) | 0.97 |
| GCSF pg/mL | 29.6(16.9-81.8) | 29.9(18-63.6) | 28.4(16.4-51.4) | 0.28 |
| **rs1800872**<br>**(-592 C/A)** | **Total** | **C** | **A** | **P**<br>**value** |
| TNFα pg/mL | 63.7(20.5-191.6) | 63.7(18.6-205.6) | 63.7(21.8-173.7) | 0.71 |
| IFN α pg/mL | 17.9(10.8-34.3) | 17.6(10.9-37.7) | 18.1(9.9-32.3) | 0.40 |
| IFN γ pg/mL | 52.9(39-91.7) | 51.4(38.3-95.5) | 53.7(40.9-84.9) | 0.91 |
| IL-1Ra pg/mL | 224.8(66.7-424.1) | 215.4(55.4-397.2) | 248.8(85.4-434.1) | 0.12 |
| IL-2 pg/mL | 27.3(16.9-81) | 27.5(16.3-85.6) | 27(17-79.3) | 0.97 |
| IL-6 pg/mL | 120.6(35.7-308.7) | 109.9(30.6-329.9) | 126.6(43.6-304.4) | 0.45 |
| IL-7 pg/mL | 128.4(73.9-436.8) | 130.9(75.17-487.9) | 123.2(70.32-395.8) | 0.57 |
| IL-10 pg/mL | 40.6(20.12-102.5) | 40.6(19.9-109.3) | 39.04(20.3-95.2) | 0.32 |
| CCL2 pg/mL | 1214.3(415.1-2470.8) | 1162.1(342.9-2470.8) | 1320.8(506.7-2470.8) | 0.16 |
| CCL3 pg/mL | 180.9(30.8-375.8) | 168.3(17.9-363.5) | 209.7(46.7-386.8) | 0.20 |
| CXCL8 pg/mL | 189.9(62.8-343.1) | 189.9(57-365.1) | 192.9(73.2-309.8) | 0.98 |
| CXCL10<br>pg/mL | 1329.7(667.1-220.6) | 1348.9(667.1-22020.6) | 1317.2(666.3-2220.6) | 0.94 |
| GCSF pg/mL | 29.6(16.7-58.5) | 29.7(17-64.2) | 28.1(16.4-51.2) | 0.28 |
| **CXCL6** |  |  |  |  |
| **rs4279174** | **Total** | **G** | **C** | **P**<br>**value** |
| TNFα pg/mL | 57.5(19.7-174.7) | 53.5(18.6-172.1) | 72.1(22.9-192.8) | 0.25 |
| IFN α pg/mL | 17.8(10.7-32.6) | 17.2(10.1-31.7) | 18.5(11.7-41.2) | 0.18 |
| IFN γ pg/mL | 50.8(37.8-88.1) | 48.7(37.4-84.6) | 55.8(40.2-95.2) | 0.20 |
| IL-1Ra pg/mL | 215.4(50.4-401.5) | 215.4(49.62-397.2) | 215.4(61.6-405.7) | 0.91 |
| IL-2 pg/mL | 25.9(16.6-77.6) | 25.8(16.2-76.7) | 26.7(18.5-79.3) | 0.45 |
| IL-6 pg/mL | 109.9(31.2-298.5) | 106.8(31.8-304.4) | 119.6(30.6-273.5) | 0.93 |
| IL-7 pg/mL | 119.2(72.1-403.3) | 116.4(65.5-404.8) | 132.6(82.9-352.8) | 0.30 |
| IL-10 pg/mL | 39.5(20.3-101.3) | 38.6(20.2-101.5) | 40.4(23.1-97.4) | 0.74 |
| CCL2 pg/mL | 1161.2(346.6-2350.5) | 1147.6(342.9-2374.9) | 1260.26(361.1-2169.9) | 0.70 |
| CCL3 pg/mL | 166.9(24.89-362.07) | 159.1(23.13-352.82) | 237.71(32.4-372.11) | 0.29 |
| CXCL8 pg/mL | 177.02(52.99-323.64) | 175.46(51.29-338.2) | 178.6(57.7-312.4) | 0.89 |
| CXCL10 pg/mL | 1278.1(647.0-2213.7) | 1237.4(629.9-2151.1) | 1395.8(667.0-2360.9) | 0.23 |
| GCSF pg/mL | 29.2(17.2-54.9) | 29.6(16.9-58.5) | 25.5(17.9-52) | 0.18 |
| **CCL5** |  |  |  |  |
| **rs2107538** | **Total** | **C** | **T** | **P value** |
| TNFα pg/mL | 58.8(19.5-179.9) | 51.6(19.5-168.7) | 85.7(22.2-205.6) | 0.09 |
| IFN α pg/mL | 17.6(10.62-32.91) | 16.68(10.1-30.85) | 19.68(11.84-35.06) | 0.05 |
| IFN γ pg/mL | 51.1(38.3-90.9) | 48.9(37.8-87.3) | 55.8(40.9-95.5) | 0.16 |
| IL-1Ra pg/mL | 215.7(61.3-424.1) | 201.5(42.2-424.1) | 230.4(95.6-408.4) | 0.11 |
| IL-2 pg/mL | 26.8(16.9-78.7) | 24.3(15.9-67.9) | 33.3(18.5-95.5) | **0.002** |
| IL-6 pg/mL | 122.8(31.8-306.6) | 97.7(29.9-273.6) | 146.7(44.4-358.4) | **0.02** |
| IL-7 pg/mL | 125(73.6-404.8) | 112.5(64.5-337.7) | 167.5(81-517.9) | **0.003** |
| IL-10 pg/mL | 39.5(20.2-101.5) | 36.2(19.8-90.9) | 54.9(24.3-131.9) | **0.02** |
| CCL2 pg/mL | 1191.66(361.49-2452.61) | 1047.31(334.44-2297.7) | 1446.3(665.68-2644.46) | **0.007** |
| CCL3 pg/mL | 166.9(24.9-372.1) | 149.8(17.9-372.1) | 220.5(37.0-380.6) | 0.14 |
| CXCL8 pg/mL | 179.9(57.4-325.1) | 171.3(42.6-286.3) | 206.7(103.3-382.1) | **0.002** |
| CXCL10 pg/mL | 1310.7(666.3-2206.7) | 1182.5(605.3-2117.9) | 1611.5(697.1-2652.8) | **0.02** |
| GCSF pg/mL | 29.2(16.5-54.5) | 27.3(16.3-51.2) | 36.7(20.5-64.2) | **0.02** |
| **rs3817656** | **Total** | **A** | **G** | **P value** |
| TNFα pg/mL | 54.4(18.3-173.7) | 51.2(18.6-168.7) | 75.6(17.4-191.6) | 0.37 |
| IFN $\alpha$ pg/mL | 17.6(10.2-31.9) | 17.0(10.1-30.8) | 19.7(10.79-32.9) | 0.26 |
| IFN $\gamma$ pg/mL | 48.9(37.4-84.9) | 48.7(37.4-84.2) | 54.4(37.4-93.1) | 0.45 |
| IL-1Ra pg/mL | 215.3(51.1-405.8) | 199.5(42.2-376.0) | 230.4(85.3-408.4) | 0.11 |
| IL-2 pg/mL | 25.7(16.5-70.9) | 24.0(15.5-64.2) | 30.9(17.9-92.2) | **0.008** |
| IL-6 pg/mL | 109.9(31.8-306.6) | 95.7(29.5-254.8) | 137.2(44.0-400.7) | **0.02** |
| IL-7 pg/mL | 116.4(70.3-352.8) | 111.2(64.5-263.8) | 133.35(79.94-484.67) | **0.03** |
| IL-10 pg/mL | 37.6(20.1-97.4) | 35.4(19.8-83.9) | 50.9(23.1-114.9) | 0.11 |
| CCL2 pg/mL | 1162.0(352.8-2374.9) | 1032.9(327.4-2283.8) | 1425.7(665.7-2653.5) | **0.02** |
| CCL3 pg/mL | 164.7(23.1-368.3) | 153.8(7.6-368.3) | 215.1(37.0-352.8) | 0.36 |
| CXCL8 pg/mL | 178.6(52.3-311.5) | 171.3(41.8-284.5) | 200.7(97.0-366.6) | **0.01** |
| CXCL10 pg/mL | 1299.9(642.4-2206.7) | 1182.0(601.8-2124.4) | 1545.3(666.3-2567.9) | 0.14 |
| GCSF pg/mL | 28.2(16.7-54.5) | 26.4(46.4-51.4) | 31.4(18.3-63.0) | 0.12 |
| **IL-10 HAPLOTYPES** |  |  |  |  |
| TNFα pg/mL | 67.8(20.8-192.8) | 67.8(19.5-205.6) | 67.8(22.4-179.9) | 0.82 |
| IFN $\alpha$ pg/mL | 18(10.8-35.01) | 17.7(10.9-37.6) | 18.1(9.8-33.3) | 0.45 |
| IFN $\gamma$ pg/mL | 53.9(39.0-93.1) | 51.7(38.5-96.2) | 54.7(40.9-85.9) | 0.94 |
| IL-1Ra pg/mL | 228.9(70.5-434.1) | 215.9(61.3-420.6) | 255.1(105.3-442.2) | 0.08 |
| IL-2 pg/mL | 27.8(16.9-85.5) | 27.8(16.7-87.5) | 27.4(17.6-81.8) | 0.92 |
| IL-6 pg/mL | 125.7(37.6-307.7) | 114.3(31.8-329.9) | 126.8(45.9-304.4) | 0.39 |
| IL-7 pg/mL | 130.3(76.4-466.3) | 132.9(76.4-488.1) | 130.2(76.4-401.9) | 0.65 |
| IL-10 pg/mL | 41.1(20.1-102.5) | 40.6(19.9-109.3) | 42.8(20.2-95.2) | 0.37 |
| CCL2 pg/mL | 1277.3(439.2-2506.2) | 1172.9(345.5-2506.2) | 1324.8(525.9-2644.5) | 0.14 |
| CCL3 pg/mL | 191.3(33.1-380.6) | 173.4(23.1-363.5) | 221.2(54.9-387.2) | 0.11 |
| CXCL8 pg/mL | 195.6(67.2-344.3) | 190.1(57.7-365.1) | 197.8(78.9-312.4) | 0.75 |
| CXCL10 pg/mL | 1336.8(670.5-220.6) | 1378.3(670.5-2220.6) | 1312.0(670.5-2220.6) | 0.99 |
| GCSF pg/mL | 29.6(16.9-60.03) | 30.01(17.9-64.1) | 28.8(16.4-51.4) | 0.34 |
| **CCL-5 HAPLOTYPES** |  |  |  |  |
| TNFα pg/mL | 54.5(18.6-173.7) | 51.2(18.6-168.7) | 75.6(20.8-191.6) | 0.23 |
| IFN $\alpha$ pg/mL | 17.5(9.9-31.8) | 16.1(9.7-27.9) | 19.7(10.8-32.9) | 0.11 |
| IFN $\gamma$ pg/mL | 48.9(37.4-87.4) | 48.7(37-84.3) | 54.6(37.8-93.1) | 0.31 |
| IL-1Ra pg/mL | 215.4(61.3-434.1) | 201.5(44.8-433.2) | 250.3(101.7-440.5) | 0.07 |
| IL-2 pg/mL | 25.8(16.8-72.8) | 24.2(15.9-64.0) | 32.5(18.3-92.2) | **0.003** |
| IL-6 pg/mL | 120.06(31.9-323.4) | 101.3(29.9-294.3) | 142.7(44.4-400.6) | **0.02** |
| IL-7 pg/mL | 116.4(70.6-360.5) | 111.2(64.5-263.8) | 133.5(79.9-488.1) | **0.01** |
| IL-10 pg/mL | 37.4(19.9-97.4) | 34.9(19.5-78.4) | 50.9(23.1-117.4) | 0.05 |
| CCL2 pg/mL | 1191.7(376.9-2506.2) | 1078.8(342.9-2374.9) | 1446.3(713.8-2796.1) | **0.008** |
| CCL3 pg/mL | 164.7(24.3-380.6) | 153.8(7.6-375.8) | 218.4(37.0-386.8) | 0.22 |
| CXCL8 pg/mL | 179.9(57.4-309.8) | 173.2(48.8-284.5) | 206.7(103.7-366.6) | **0.005** |
| CXCL10 pg/mL | 1309.5(657.2-2131.3) | 1182.1(605.3-2118.4) | 1545.3(666.3-2626.5) | 0.07 |
| GCSF pg/mL | 28.2(16.4-53.3) | 26.4(15.7-51.2) | 33.7(18.1-64.1) | 0.05 |

8 P-value obtained from Kruskal-Wallis test. Values in bold denotes statistical significance

Interestingly, we observed significant differences for the rs1049953 (−6331T/C) polymorphism of the *IL-6* gene with respect to the level of *IFN-*α. In addition, we observed that the carriers of the risk allele of this SNP had the highest serum concentrations of IL-6 and CCL-2 (Table 4).

Similarly, for the *CCL-5* rs2107538 polymorphism, carriers of the A allele had the highest serum levels of the IL-2, IL-6, IL-7, IL-10, CCL-2, GCSF, and CXCL10 cytokines. For rs3817656, carriers of the risk allele presented increased serum levels of the cytokines IL-2, IL-6, IL-7, CCL-2, GCSF, and CXCL8. These differences were also found to be significant for the *CCL-5* haplotype AG, with the exception of the GCSF concentration, which did not reach significance (Table 4).

We performed a Spearman correlation analysis with the WHO severity score and observed a moderate positive correlation with IL-6 (ρ=0.53), IL-1 (ρ=0.52) and CCL2 (ρ=0.43) (p≤0.0001).

## Discussion

COVID-19 inflammation has been described; however, it is still actively being studied. The inflammatory process is characterized by a cytokine storm. Severe COVID-19 patients have increased levels of proinflammatory and anti-inflammatory cytokines and chemokines, such as IL-6, TNF-α, IL-10, CCL2, CCL3, CXCL8, and CXCL10, among others (25–28). Since COVID-19 emerged, several studies across the world have reported some polymorphisms of different genes related to this disease (29). Understanding the host genetic basis of inflammation could help clarify the key mechanisms through which patients experience worsening of COVID-19 severity. In the present study, we analyzed SNPs in the *TNF-*α*, IL-6, IL-8*, *IL-10*, *CCL5* and *CXCL6* genes associated with COVID-19 outcomes and with serum cytokine and chemokine levels.

Compared with other diseases, such as influenza A virus, severe acute respiratory syndrome (SARS) and Middle East respiratory syndrome (MERS), COVID-19 is characterized by greater dysregulation of inflammatory molecules, which may contribute to the severity of the disease (25, 30). In the present study, we observed a trend toward increased CCL2 levels among COVID-19 patients, with the levels of IL-Ra and CCL3 being elevated in patients with severe disease. These findings are similar to those reported by Torres-Ruiz et al. (2021), who reported a tendency for CCL2, CCL3 and other cytokines and chemokines to increase with disease severity (30). García-Ramírez et al. (2015) proposed that inflammatory cytokines play crucial roles in the progression of diseases caused by different types of infections. Additionally, a study by Dorgham et al. (2021) revealed increased levels of IFN-α, TNF-α and IL-10 in patients who died from COVID-19, suggesting that the cytokine profile may have clinical implications and could help in the personalized management of this disease (31–34). Interestingly, we observed higher concentrations of these cytokines in patients with moderate outcomes.

According to previous studies that revealed increased levels of pro- and anti-inflammatory cytokines, genetic variation could be involved in the host inflammatory response, affecting the severity of COVID-19. Several genome-wide association studies (GWASs) and candidate gene studies have identified SNPs associated with susceptibility to SARS-CoV-2 infection and severe COVID-19 (5, 6, 34, 35). In this sense, we studied the genetic variants of the *TNF*-α (rs1800610, rs1800629 (−308 G/A), rs3093664), *IL-6* (rs1800796 (−572 G/C), rs10499563 (−6331 T/C)), *IL-8* (rs2227307), *IL-10* (rs1800872 (−819 C/T) and rs1800071 (−592 C/A)), *CXCL6* (rs4279174), and *CCL5* (rs2107538, rs3817656) genes.

In COVID-19, TNF-α-mediated inflammation promotes detrimental tissue damage and gradual lung fibrosis, which results in pneumonia, pulmonary edema, and acute respiratory distress syndrome. These conditions are considered independent risk factors for death among COVID-19 patients with critical conditions (26). In addition, the *TNF-*α gene is highly polymorphic, and the most studied variants are rs1800610, which is located in the first intron of the gene +489 (G/A), and rs1800629, which is located in the promoter region of the gene −308 (G/A). However, its role during COVID-19 infection is still controversial. Our results revealed that only the rs1800610 A allele was significantly associated with the risk of a moderate COVID-19 outcome. The rs1800610 A allele in the Australian population was associated with a greater risk of developing breathing problems with bronchial exacerbation, suggesting that this SNP could contribute to disease progression (36). In addition, rs1800610 tends to increase cytokine concentrations in carriers of the A allele, and this SNP is related to prostate cancer and COPD development(36, 37). rs1800629 (−308 G/A) was found to be associated with protection against moderate and severe disease. The protective effect of rs1800629 (−308 G/A) is consistent with that reported by Heideri Nia et al., who showed that allele A decreases the risk of SARS CoV 2 infection (38).

The role of IL-6 in COVID-19 has been described, suggesting that the IL-6 family can regulate genes and proteins involved in angiogenesis and immune cell recruitment (39). However, IL-6 has a pleiotropic effect on patients that is age dependent (13). Our models of associations were adjusted for principal comorbidities such as age, sex, type 2 diabetes, obesity and hypertension.

The *IL-6* gene variants rs1800796 (−572 G/C) and rs10499563 (−6331 T/C), which are located in the promoter region, are associated with immunological regulation of hepatocellular carcinoma (40, 41) and other inflammatory pathologies, such as gastric cancer (42), steatohepatitis (43), hepatitis B (44) and frailty (45). The rs1800796 (−572 G/C) SNP contributes to the functional regulation of expression of this gene (46).

A study by Rodrigues *et al.* reported an association between the rs1800795 polymorphism in the *IL-6* promoter region and COVID-19 severity. They found that plasma IL-6 levels were greater in carriers of the risk allele. While the SNP examined in their study was different from that investigated in the present work (47), we observed similar results. Specifically, we found that the C allele of the (−572 G/C) rs1800796 polymorphism was associated with severe COVID-19. These findings are in agreement with previous studies that linked *IL-6* SNPs to the clinical progression of COVID-19 (48). However, Falahi *et al*. reported no association between the (−572 G/C) rs1800796 polymorphism and COVID-19 severity in the Kurdish population (49). Furthermore, Karcioglús study, which explored the correlation between rs1800796 (−572 G/C) frequency and COVID-19 incidence and mortality rates across 23 countries, revealed no significant correlation. These discrepancies could be attributed to variations in the genetic background of the populations studied. Nevertheless, the authors suggested that SNPs in *IL-6* could be involved in the regulation of its expression (50). In our study, we demonstrated that rs1800796 (−572 G/C) of the *IL-6* gene was associated with severe COVID-19 in the Mexican population.

IL-8 has effects through binding to cognate G-protein-coupled CXC chemokine receptors, CXCR1 and CXCR2, which activate a phosphorylation cascade to trigger chemotaxis and neutrophil activation as part of the inflammatory response. In COVID-19 patients, the plasma IL-8 concentration is increased, suggesting a significant role in cytokine release syndrome, multiorgan dysfunction, respiratory failure and shock (51, 52). SNPs in the *IL-8* gene could influence the molecular mechanisms of COVID-19. Nevertheless, in the present study, we did not find an association between rs2227307 and COVID-19 severity. The *IL-10* gene is located on chromosome one and has five exons and four introns. The SNPs rs1800871 (−819 C/T) and rs1800872 (−592 C/A), located in the 5’-flanking region, are involved in the regulation of IL-10 production (53). In autoimmune diseases and ankylosing spondylitis (AS), increased levels of IL-10 in the serum have been described.

Huang et al. (2020) reported that IL-10 serum levels are greater in patients admitted to the ICU than in those who are not (34). IL-10 is produced by type 2 T helper cells and type 2 macrophages, and elevated levels of this cytokine are associated with endothelial dysfunction. In addition, in the *IL-10* gene, for the polymorphism rs1800871 (−819 C/T), the allele T was found in more than 50% of the population, and CT was found in 48%. Additionally, in r1800872 (−592 C/A), the CA genotype was observed in 54% of the deceased individuals, and this polymorphism could be considered a susceptibility factor in our population study. Both polymorphisms were associated with severe and deceased COVID-19 outcomes. Previously, in Mexico, rs1800871 (−819 C/T) and r1800872 (−592 C/A) were found to not be associated with COVID-19 severity. However, the sample size was small (193 subjects), which means that the power was low (54). Abbood et al. (2023), in an Iranian population, reported that rs1800871 (−819 C/T) and r1800872 (−592 C/A) were related to COVID-19 mortality(55). It has been reported that r1800872 (−592 C/A) can reduce negative promoter function, modify IL-10 transcription and play an important role in diseases such as gastric cancer, lung cancer and cervical cancer (1, 56, 57). Monroy-Muñiz et al. (2023) studied rs1800871 (−819 C/T) and r1800872 (−592 C/A) in a Mexican population and reported that the T and A alleles were associated with the risk of dengue infection (58).

CXCL6 is a chemokine characterized by four conserved cysteine residues close to the amino terminus CXC motif; this chemokine has been associated with lung cancer, gastric cancer and cervical cancer. In lung inflammation, CXCL6 stimulates inflammation by recruiting and activating immune cells. In the present study, we did not observe a significant association between rs4279174 of the *CXCL6* gene and COVID-19 severity. However, other variants in this gene may influence disease severity.

CCL5, also called RANTES, has been studied in many diseases, such as inflammation, cancer, and virus infection. Most inflammatory cells can express CCL5; recently, the CCL5/CCR5 axis was reported to participate in the response to COVID-19. Additionally, the CCL5/CCR5 axis is related to HIV and tuberculosis. Kouhpayeh et al. reported that the GA genotype in the dominant model of *CCL5* (rs2107538) was associated with an increased risk of pulmonary tuberculosis (59). Our results revealed that this SNP in the recessive model was associated with COVID-19 severity. However, Pati et al reported that this variant was associated with protection against SARS-CoV-2 infection, but the mechanisms are unknown and were not controlled by confounding factors such as sex or age, among others (60). In the present study, we found a significant association of the risk alleles of rs2107538 and rs3817656 with the severity of COVID-19, and after adjusting for sex, age, type 2 diabetes and hypertension, we also observed higher concentrations of cytokines in the presence of the risk alleles. The rs21075238 is located at the binding site for GATA binding protein 2 (GATA2), which is involved in the transcriptional regulation of proinflammatory cytokines, and CCL5 might participate in the COVID-19 inflammatory process (61–63).

Higher levels of diverse cytokines and CXCs have been described to be associated with COVID-19 severity (64). CXCs constitute a subfamily of chemotactic cytokines, and 17 CXC chemokines have been described; this subfamily has also been associated with tumors and inflammatory diseases (16, 65). In COVID-19, CXCL9 and CXCL10 have been implicated in respiratory failure and death (65).

CCL-2 is an important mediator in multiple lung inflammatory diseases, and its major mechanism involves mediating the migration of mononuclear cells into the microvasculature and airways. This could explain the monocyte and lymphocyte influx observed in ARDS, asthma and bronchiolitis obliterans syndrome (BOS) (66, 67).

A recent report by Abers *et al.* suggested that CCL-2 was associated with mortality due to COVID-19 (68). Pius-Sadowska *et al.* reported that lower concentrations of CCL-2 mRNA but higher concentrations of the protein in plasma were correlated with more severe courses of COVID-19, suggesting that CCL-2, like other cytokines, could be considered new prognostic factors for severe COVID-19 (69).

When we explored the cytokine serum concentrations stratified by alleles of the evaluated SNPs, we observed that the rs1800629 A allele polymorphism was associated with the lowest levels of IL-1Ra, IL-6, CCL2 and CXCL10. We suggest that this SNP could have downstream effects on IL-1Ra, IL-6, CCL2 and CXCL10, which are involved in COVID-19.

Additionally, we observed increased levels of IFN-α, IL-6 and CCL2 in patients with the variant alleles of the rs10499563 (−6331 T/C), rs2107538, rs3817656 and CCL5 haplotypes compared with the ancestral allele. These SNPs could contribute to increased IL-6 levels, which can increase CCL2 production and promote immune cell recruitment. While the impact of these SNPs on IFN-α is less clear, IL-6 can influence the broader immune environment, potentially enhancing IFN-α responses in viral infections (10, 16, 34, 35). Previous studies reported that plasma levels of IL-6 were associated with worse clinical outcomes in patients with SARS-CoV-2(70).

This is the first work to evaluate the genetic profile and cytokine levels associated with COVID-19 severity in a nonvaccinated Mexican population, adjusted for main comorbidities such as age, sex, type 2 diabetes, obesity and hypertension. However, this study has several limitations, such as the fact that patients with other pathologies could have an impaired immune system.

Further investigations of other SNPs in these genes or others implicated in the mechanism of COVID-19 severity are necessary for developing prevention actions and precision medical therapies. It should be noted that our results are only representative of the Mexican-Mestizo population.

This study highlights the associations of the *IL-6* rs1800796 (−572 G/C), *IL-10* rs1800871 (−819 C/T), rs1800872 (−592 C/A) and *CCL5* rs2107538 polymorphisms with fatal COVID-19 outcomes and rs1800629 of TNF*-*α gene with protection to moderate outcome. In this sense, the elevation of cytokines (CCL-2, IL-6, IL-10 and CXC-L10) plays a crucial role in the pathogenesis of COVID-19, contributing to multiorgan dysfunction. The *CCL5* SNPs rs10499563 (−6331 T/C), rs2107538, and rs3817656 may influence IL-6 levels, which can indirectly affect the production of CCL2 and IFN-α and could contribute to the severity of COVID-19.

## Acknowledgments

We are grateful for the extraordinary efforts of health care workers, who sacrificed their lives while saving patients. This study was funded by the Consejo Nacional de Ciencia y Tecnología; CONACYT 312513 SARS-COV 2.

## Data Availability Statement

The datasets presented in the current study are available on request from the corresponding author. The data are not public available due to privacy or ethical restrictions.

## Author Contributions

Laura E. Martínez-Gómez, Methodology, Formal analysis, Writing original draft, Writing-review and editing; Carla Isabel Oropeza-Vélez Investigation, Methodology, Writing-review and editing; Maylin Almonte-Becerril Investigation, Methodology, Writing-review and editing; Leslie Chavez-Galan Resources, Methodology, Writing-review and editing; Carlos Martinez-Armenta Resources, Writing-review and editing; Rosa P. Vidal-Vázquez Investigation, Methodology, Writing-review and editing; Juan P. Ramírez-Hinojosa Methodology, Writing-review and editing; Paola Vázquez-Cárdenas Visualization, Writing-review and editing; Diana Gómez-Martín Visualization, Writing-review and editing; Gilberto Vargas-Alarcón Visualization, Writing-review and editing; José M. Rodríguez-Pérez Visualization, Writing-review and editing; Lucero A Ramón-Luing Formal analysis, Writing-review and editing; Julio Flores-Gonzalez Formal analysis, Writing-review and editing; José G. Carrasco Writing-review and editing; Mónica M. Mata-Miranda Investigation, Writing-review and editing; Gustavo J. Vázquez-Zapién Data curation, Writing-review and editing; Adriana Martínez-Cuazitl Methodology, Writing-review and editing; Parra-Torres NM Data curation, Writing-review and editing; Felipe de J. Martínez-Ruiz Data curation, Writing-review and editing; Dulce M. Zayago-Angeles Data curation, Writing-review and editing; Ma. Luisa Ordoñez-Sánchez Formal analysis, Writing-review and editing; Yayoi Segura-Kato Data curation, Writing-review and editing; Carlos Suarez-Ahedo Methodology, Writing-review and editing; Jessel Olea-Torres Methodology, Writing-review and editing; Brígida Herrera-López Methodology, Writing-review and editing; Carlos Pineda Supervision Writing-review and editing; Gabriela A. Martínez-Nava Validation, Supervision, Writing-review and editing; Alberto G. López-Reyes Conceptualization, Project administration, Resources, Supervision, Writing-review and editing.

